# Global surveillance of potential antiviral drug resistance in SARS-CoV-2: proof of concept focussing on the RNA-dependent RNA polymerase

**DOI:** 10.1101/2020.12.28.20248663

**Authors:** Alfredo Mari, Tim Roloff, Madlen Stange, Kirstine K. Søgaard, Erblin Asllanaj, Gerardo Tauriello, Leila Tamara Alexander, Michael Schweitzer, Karoline Leuzinger, Alexander Gensch, Aurélien Martinez, Julia Bielicki, Hans Pargger, Martin Siegemund, Christian H. Nickel, Roland Bingisser, Michael Osthoff, Stefano Bassetti, Parham Sendi, Manuel Battegay, Catia Marzolini, Helena M.B. Seth-Smith, Torsten Schwede, Hans H. Hirsch, Adrian Egli

## Abstract

Antiviral treatments for COVID-19 have involved many repurposed drugs. Currently, SARS-CoV-2 RNA-dependent RNA polymerase (RdRp, encoded by *nsp12-nsp7-nsp8*) has been targeted by numerous inhibitors with debated clinical impact. Among these, remdesivir has been conditionally approved for the treatment of COVID-19 patients. Although the emergence of antiviral resistance, an indirect proxy for antiviral efficacy, poses a considerable healthcare threat, an evolutionary perspective on emerging resistant mutants is still lacking.

Here we show that SARS-CoV-2 RdRp is under purifying selection, that potential escape mutations are rare, and unlikely to lead to viral fitness loss.

In more than 56,000 viral genomes from 105 countries dating from December 2019 to July 2020 we found negative selective pressure affecting *nsp12* (Tajima’s D = −2.62), with potential antiviral escape mutations in only 0.3% of sequenced genomes. Those affected known key residues, such as Nsp12:Val473 and Nsp12:Arg555. Of the potential escape mutations found globally, *in silico* structural models show that this rarely implies loss of stability in RdRp. No potential escape mutation were found in our local cohort of remdesivir treated patients from the first wave (n=8). Our results indicate that RdRp is a suitable drug target, and that remdesivir does not seem to exert high selective pressure. Our study could be the starting point of a larger monitoring effort of drug resistance throughout the COVID-19 pandemic. We recommend the application of repetitive genome sequencing of SARS-CoV-2 from patients treated with antivirals to provide early insights into the evolution or antiviral resistance.

## Introduction

Infection with SARS-CoV-2 is associated with substantial morbidity and mortality. As no approved therapy is available to-date, there have been recently multiple efforts in drug repurposing. The selective pressure on the virus generated by potential antiviral drugs and the eventual emergence of antiviral resistance provides interesting information about the mode of action of antiviral drugs and their efficacy. In this context, the surveillance of resistance emergence is paramount for public health during the COVID-19 pandemic. The first drugs considered for antiviral treatment were inhibitors targeting 3C-like proteases (3CL-Pro) and Spike proteins (S)^1,2^. However, the alarming number of side effects and the lack of clinical efficacy forced the identification of new targets of viral replication machinery^3^. Because of its high degree of aminoacid conservation within beta-coronaviridae, RNA dependent RNA polymerase (RdRp) (96% identity^4^) is a key target of antiviral drug development, and recently, of drug-repurposing^5,4^. Several studies have indicated *in vitro* efficacy against SARS-CoV-2 of potential inhibitory candidates such as remdesvir, sofosbuvir, galidesivir, tenofovir, and ribavirin^6,7,8^. All of them share the same mechanism of action, binding RdRp in its active site as nucleoside analogues, interrupting the RNA polymerisation^6^ **(Figure S1)**.

Antiviral treatments have been shown to lead to the emergence of antiviral resistance in infectious diseases such as those caused by Hepatitis B Virus, Human Immunodeficiency Virus, and Hepatitis C Virus (HCV), or influenza itself^9,10,11^. For example, the emergence of ribavirin resistant mutants has been described for HCV RdRp and has been largely attributed to the emergence of resistance-conferring single nucleotide polymorphisms (SNPs), usually resulting in a remarkable fitness loss^10,12^. Similarly, *in vitro* experiments on SARS-CoV, cause of the Severe Acute Respiratory Syndrome (SARS), which is the closest related to SARS-CoV-2, show that specific SNPs within *nsp12* may alter the effectiveness of remdesivir: namely Nsp12:Phe480Leu and Nsp12:Val557Leu^13^. The substitution Nsp12:Phe480Leu destabilises the interface between different sub-domains of the protein (“palm” and “fingers”), likely affecting the proof-reading capacity of RdRp^4^. Nsp12:Val557Leu affects binding to the template RNA and indirectly to remdesivir^4^. The EC50 of remdesivir increased six-fold from 0.01µM to 0.06µM in cultures of SARS-CoV carrying Nsp12:Phe480Leu or Nsp12:Val557Leu mutations^13^. In the absence of remdesivir, these viral mutants were found to replicate less efficiently, and showed a substantially reduced fitness^13^.

The clinical efficacy of remdesivir in COVID-19 treatment has been recently debated. Two studies suggested a reduction in recovery time^14,15^, while others did not show a reduction of mortality^16,17^. At present, expert panels from authorities have made recommendations on the use of remdesivir in patients with different severities of disease. (https://www.covid19treatmentguidelines.nih.gov/whats-new/).

Identification and surveillance of potential antiviral resistant strains and transmission thereof is essential for disease surveillance. In the genomes of circulating SARS-CoV-2, no SNPs have been associated with clinical failure of remdesivir treatment to date, although mutations in *nsp12* have been reported.

In this study, we address the suitability of RdRp as a valuable drug target by evaluating the selective pressure affecting RdRp and monitoring the emergence of potential escape mutations in key drug binding motifs. We screen real world data from both a global dataset consisting of more than 56,000 genomes from 105 countries, and an inpatient longitudinal genome cohort of 197 remdesivir treated patients (189 untreated patients, eight remdesivir treated patients with follow-up time points). We show that potential escape mutations to remdesivir are rare, as RdRp is under negative selection, and that these escape mutations generally do not hamper RdRp stability, such that compensatory mutations are not necessary.

## Results

### Selection analysis of *nsp12*

Understanding selection pressure on viral genes are critical in studying potential effects of antiviral drugs. Therefore, we inferred the selection pressures on the whole *nsp12* gene (2793 nucleotides, 931 codons) applying Tajima’s D and on the individual codons within the coding sequence applying dN/dS. In a globally subsampled (14,612 sequences) dataset of SARS-CoV-2 *nsp12*, we identified 678 segregating nucleotide sites out of 2793 (24.3% variable sites). Most codons are conserved in the protein alignment translated from the nucleotide alignment except for the amino acid position 323 (nucleotide position 967-969), where proline is often substituted for leucine. We found a nucleotide diversity (pi) of 0.000389, a population nucleotide diversity (theta) of 0.0238, and a transition/transversion ratio of 1.94. Tajima’s *D* test statistic for *nsp12* was negative (D = −2.62), indicating an excess of rare alleles in this dataset. The dN/dS analysis of *nsp12* gives a mean dN/dS of 0.61. In more detail, along the gene, few codons (357) are within the credibility range for neutral selection, and most codons (574) have a dN/dS smaller than 1, indicating different degrees of purifying selection acting on the individual codons along the gene **(SI Figure 2)**. These results indicate that nsp12 is under purifying selection, implying that accumulation and fixation of mutations is evolutionarily unfavoured with deleterious mutations being eliminated from the coding sequence.

### Identification of key motifs for drug binding sites

The structure of the active site of RdRp was screened *in silico* to identify motifs likely to be involved in nucleoside binding, to affect the binding affinity of remdesivir, and to compromise the stability or the proofreading activity of RdRp^4,18,19^. Those include Nsp12:Phe480 and Nsp12:Val557 and were identified in the buried chains starting at Nsp12:Arg467 till Nsp12:Val493 and from Nsp12:Leu544 to Nsp12:Gln570, referred to as first and second potential escape motifs, for ease of nomenclature.

### Potential escape mutants in the global dataset

Building on our published genome analysis pipeline COVGAP^20^, we developed an updated version, which is able to track genetic diversification of drug target residues, among others, the mentioned potential escape motifs of RdRp, particularly in its main chain Nsp12.

58,806 high-quality publicly available SARS-CoV-2 genomes were collected between 24th December 2019 and 12th July 2020 from GISAID^21^(**SI Table1**).

Non-synonymous mutations in the *nsp12* coding sequence were found in 46,469 of the 56,806 (81.08%) viral genomes. However, most of these mutations (e.g position C14408T leading to Pro323Leu - **SI Figure 3**) were neither within the potential escape motifs nor located around key residues of the active site which includes, among others, the residues Arg555, Lys545, Asn691, Asp623^19^. Only 182/56,806 (0.32%) genomes contained a total of 85 different non-synonymous mutations within the potential escape motifs **(SI Table2, SI Table3) (Figure 1A)**, thereby being potential candidates for reduced remdesivir effectiveness. Of the 182, three genomes exhibited non-synonymous mutations affecting Nsp12:Phe480: Nsp12:Phe480Leu, Nsp12:Phe480Ser and Nsp12:Phe480Cys. The residue Nsp12:Val557 was found to be mutated in a single genome (Nsp12:Val557Glu). Additional high frequency non-synonymous SNPs included those encoding Nsp12:Asn491Ser (occurring in 34 genomes) and Nsp12:Val473Phe (19 genomes). (**Figure 1A, SI Table3**).

**Figure 1:**
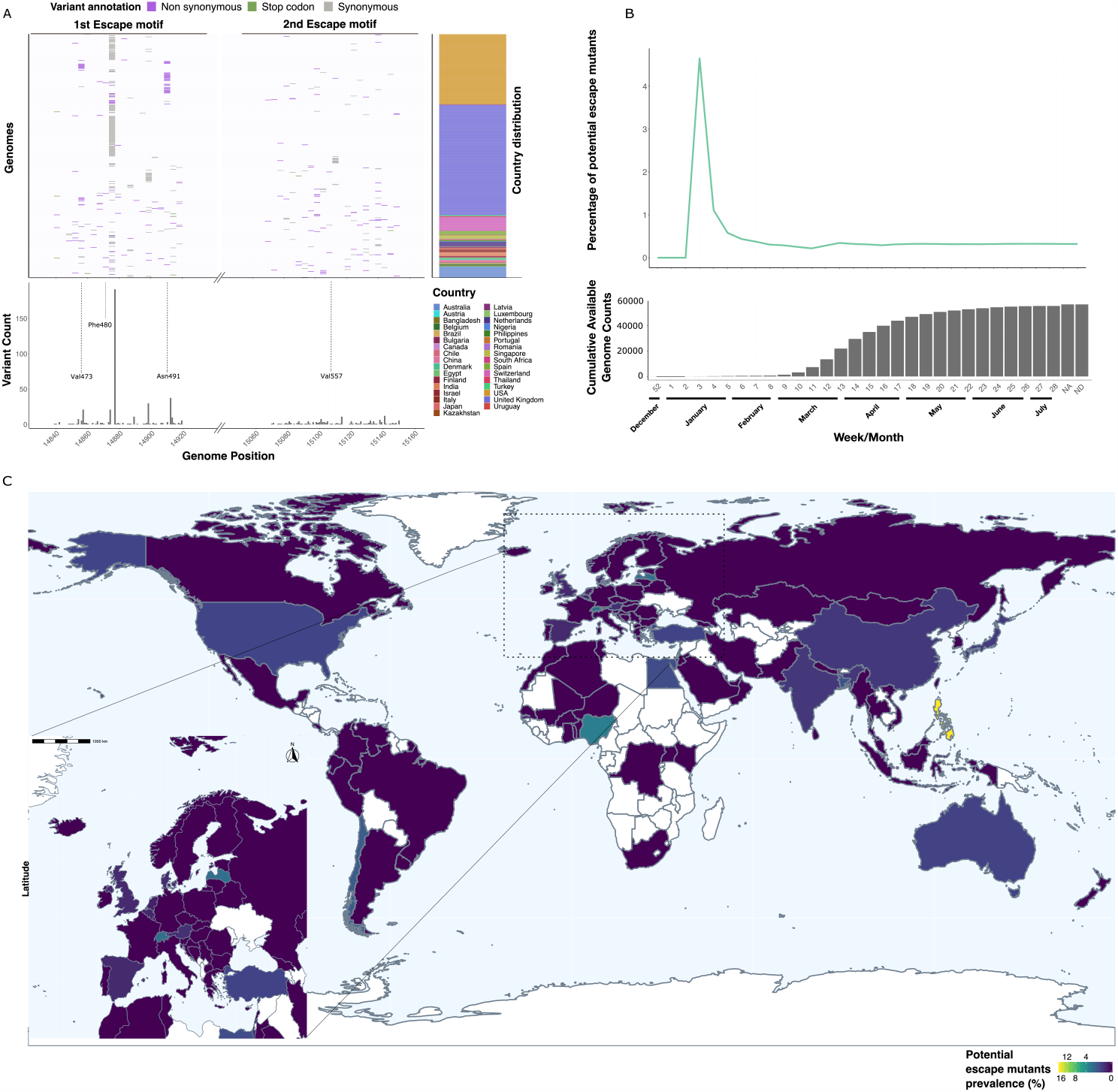
Potential escape mutations inside 1^st^ and 2^nd^ escape motif and are not uniformly distributed, they are rare worldwide and stable overtime: (A) Mutation prevalence in the two potential escape motifs, SNPs are highlighted in the upper genome panel, their frequency in the lower variant count panel, genome origin in the upper right country distribution panel, (B) Frequency changes of potentially resistant genomes over time become stable after the 13th calender week in 2020 and settle to 0.32%. The cumulative count panel displays available genomes at a defined week. The time frame considered spans from 25^th^ of December 2019 till 12^th^ of July 2020, NA/ND stand for no date information available, or incomplete respectively. (C) Escape mutants are distributed heterogeneously over the world, with Philippines harbouring 16% of potential escape mutants (total sequences n=12). In Switzerland, the potential escape mutants frequency cumulates to 2.2% (total sequences n=676), more than 7 times the global average (C-zoom).

**Figure 2:**
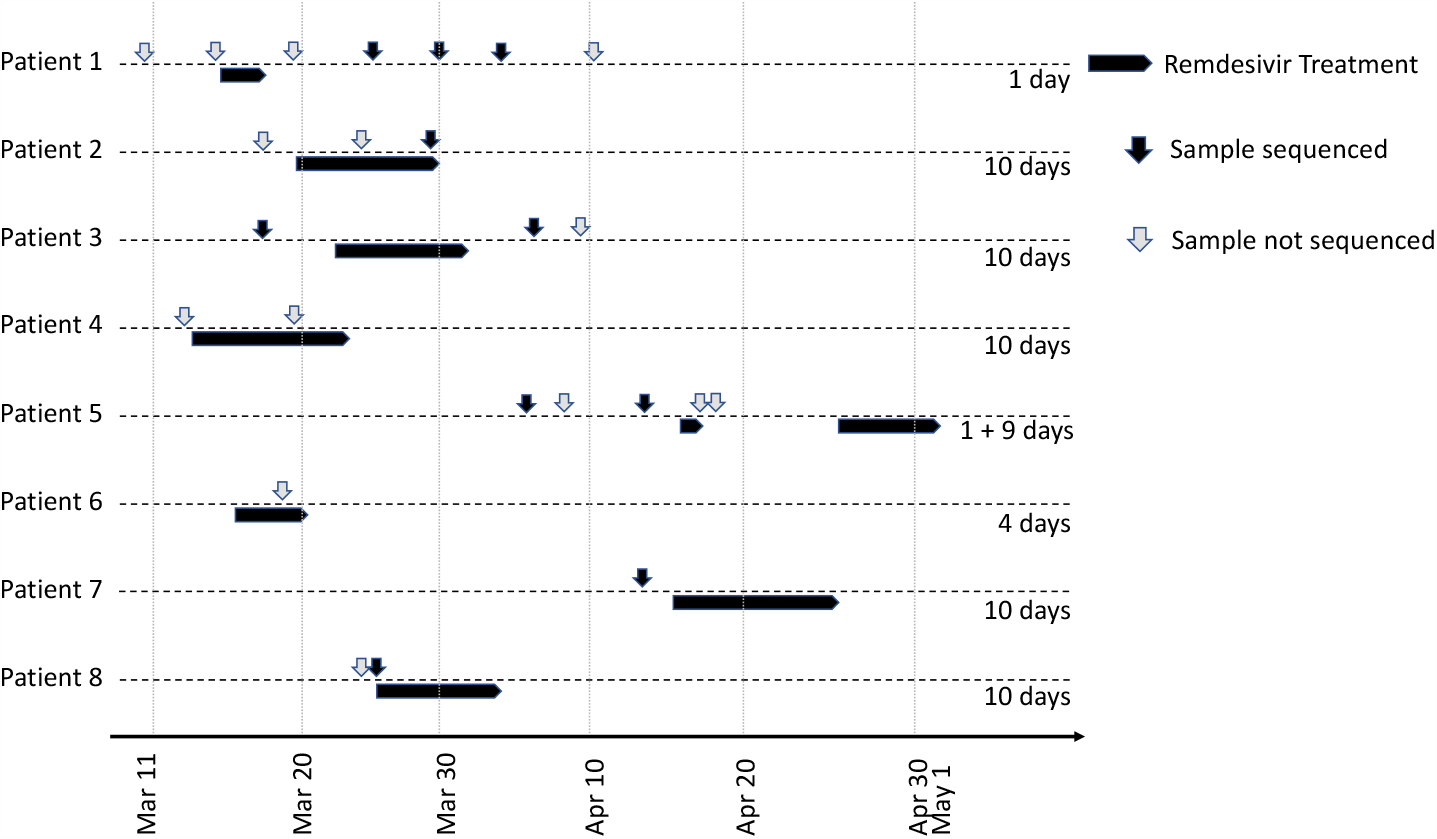
Patients who received Remdesivir treatment in the study period. Successfully sequenced samples are marked with black arrows. Grey arrows indicate samples that could not be sequenced for reasons of accessibilty, sample quality or low viral load. Remdesivir treatment is indicated by black horizontal bars.

**Figure 3:**
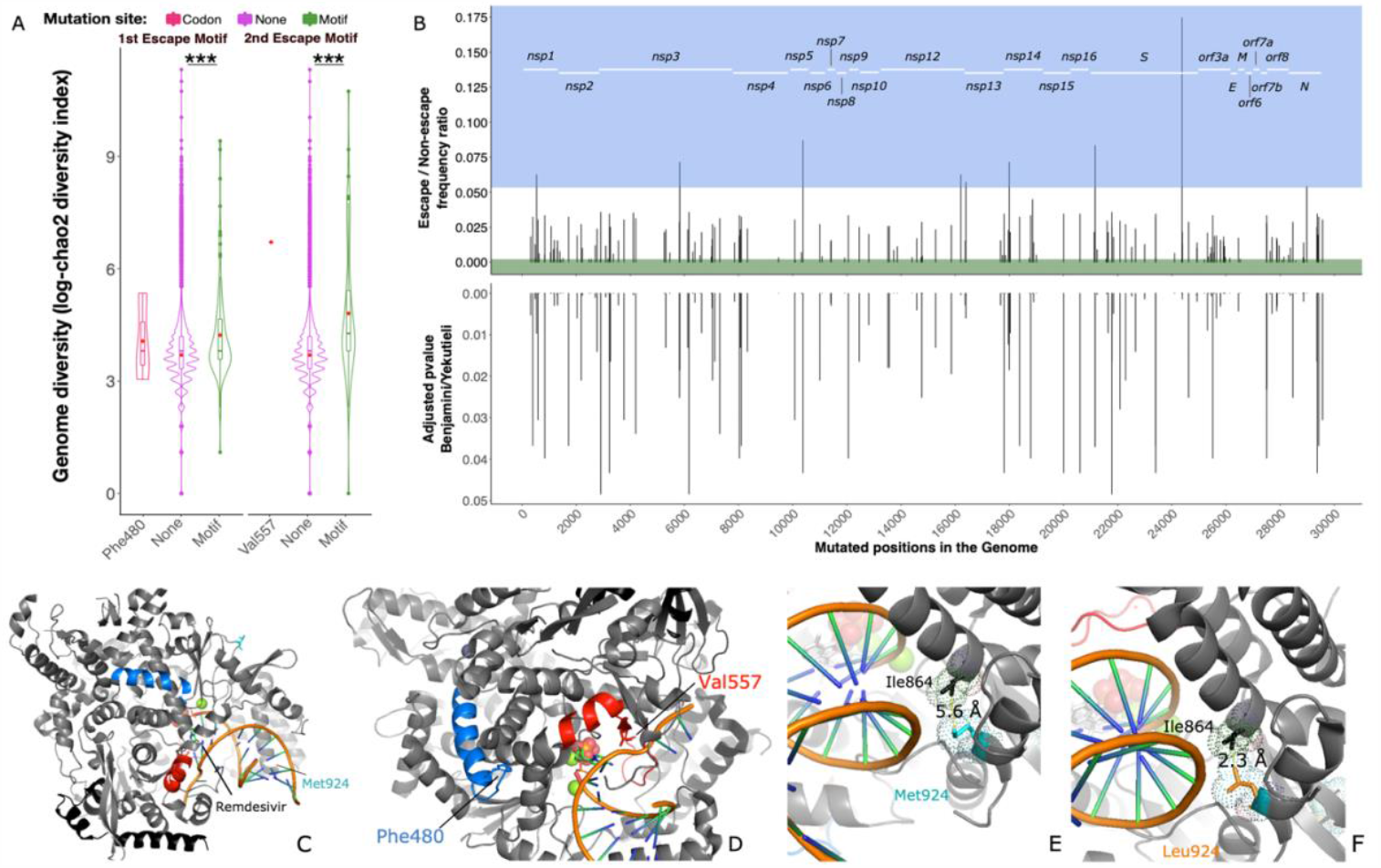
Mutations in escape motifs harbour higher genome diversity and tightly associated mutations: The genome entropy is significantly higher in 1^st^ and 2^nd^ motif escape mutants. (A) Diversity is calculated as incidence-based mutation richness along the chao2 diversity index, boxplot represents the interquartile range, red dots indicate the mean. Stars indicate the merged p-value, calculated with a Monte Carlo t-test simulation, see methods. (B) Association between escape mutants and other mutations across the genome. Candidates are evaluated through generalised linear models with lineage correction. Upper panel: depiction of mutation incidence ratio escape/non escape. Only mutations showing a ratio > 95^th^ percentile are considered escape-associated (light blue area), of note mutation in position 16,210 is significantly associated to escape mutants. Below: adjusted p-value for multiple testing according to Benjamini-Yekutieli –only mutations with significant pvalues are shown-. (C,D) Location of escape mutations and associated mutation on RdRp bound to RNA template and remdesivir, 1^st^ escape motif is indicated in blue, 2^nd^ escape motif is indicated in red. (E,F) Met924Leu (encoded by a SNP in position 16210) decreases the distance to Ile864 by more than 2-fold.

The first genome carrying a non-synonymous variant falling into a potential escape motif (Nsp12:Ser564Ile) was registered on 20th January, 2020 in a 30 year old female patient from China^22^ (**Figure 1**). The sample carrying Nsp12:Phe480Leu was collected on 3rd March 2020 from a 72 years old male patient from England. With increasing numbers of available sequences, genomes carrying non-synonymous mutations in potential escape motifs settled at 0.21% by calendar week 13, and reached a stable rate of 0.3% (+/-0.064) from week 15 on (available genomes n=35,055). No proportional increase has been detected in any time points after that date, even after conditional approval of remdesivir (**Figure 1B**). To investigate the geographical distribution of the identified escape mutants, we considered only the countries having submitted at least 100 high quality genomes. We found that Switzerland (2.2%), Chile (1.4%), and Bangladesh (1.02%) showed the highest percentages of genomes that feature potential escape mutations (**Figure 1C, SI Table 4**).

To add granularity to the Swiss data, we collected an open cohort of 690 individuals from the University Hospital Basel, who were tested only once (single-time tested) between 23rd of February and 30th of April 2020. We did not find any mutation in the potential escape motifs, nor minority alleles in the samples that would hint to intra-host diversity. The variant distribution across RdRp is in line with the distribution observed in the global dataset, with the exception of a high frequency of a synonymous mutation in nucleotide position 15324 (located in RdRp), recently described as a Basel-area specific mutation^20^ **(SI Figure 4)**.

We determined phylogenetic (pangolin nomenclature) lineages of the genomes carrying potential escape motif mutations^23^, and found that they are distributed across 37 different lineages in 21 countries. Among these is the B1.108 lineage (32 genomes), as yet only identified in the USA, first detected on 14th March 2020 and not seen after 24th April 2020. This lineage is defined by nucleotide mutation *nsp12*:A14912G, encoding a non-synonymous SNP leading to Nsp12:Asn491Ser. This lineage thus shows a potential escape mutation rate of 100% (**SI Table 5**). These results indicate that emergence of escape mutations leading to potential antiviral resistance is a rare event, independent of geographical location.

### Potential escape mutants during remdesivir treatment

From mid March 2020 on, the University Hospital Basel participated in the COVID-19 clinical trial NCT04323761 on remdesivir treatment for COVID-19 pneumonia. We collected a longitudinal cohort of 197 SARS-COV-2 positive hospitalized patients from 26th February until 30th April 2020. The samples underwent sequencing across the potential escape motifs in RdRp, bases 14545 to 15246. A total of eight patients were treated with remdesivir (duration between 2 and 10 days) **(Figure2) (SI Table 6)**. Through our COVGAP pipeline, we automatically monitored the occurrence of major and minor SNPs causing amino acid changes in any specific motifs.

We did not observe any highly supported variants in the two potential escape motifs in the remdesivir-treated patients. However, in one sample from a hospitalised 47 years old male patient, who did not receive remdesivir treatment, we observed a minority allele carrying a non-synonymous mutation in *nsp12* encoding Thr374Cys (21% alternative allele support of 27 read coverage). This substitution is not within the escape motifs and is outside the active site of RdRp. These findings are in line with the rarity of escape mutants observed in the global dataset, which support the hypothesis of a low selective pressure provided by remdesivir treatment.

### Associated mutations and stability loss

We found that the level of genetic diversity was significantly higher for potential escape mutants compared to non-mutants for both escape motifs after t-test run over Monte-Carlo simulation, used to correct for group size disparity (chi-sq =115989.4, df=20000, merged-pval << 0.001) and (chi-sq =191521.7, df=20000, merged-pval << 0.001), **(Figure 3 A)** (Chao2 diversity incidence index, **SI Table 1**).

We then screened the escape mutants to find escape-associated mutations. Within the 177 escape mutants we discarded five genomes because retaining stop-codon gain substitutions and we identified 1,056 non-synonymous potential escape-associated mutations. We evaluated lineage corrected co-occurrences through generalised linear models, and 174 non-synonymous mutations showed significant response to the predictor **(Figure 3B, SI Table 7)**. Among these, nine were positively associated with potential escape mutants.

Only one of the found escape-associated mutation falls into *nsp12*, in genome position 16210 (Nsp12:Met924Leu) **(SI Table 8)**. This variant occurs in genomes carrying the potential escape mutations Nsp12:Arg555Pro, Nsp12:Met566Val, Nsp12:Thr567Ser and Nsp12:Arg569Gly and could hamper the binding to the RNA template^19^. We found that the substitution to Leu924 shortens the distance with the close Ile864 residue from 5.6Å to 2.3Å **(Figure 3 E,F)**.

To determine the protein stability change in each scenario, we calculated the energy state of RdRp under escape and escape-associated mutation combination co-occurring in the same genomes **(SI Table9)**. We included the combinations between potential escape mutations Nsp12:Met566Val, Nsp12:Arg555Pro, Nsp12:Thr567Ser, Nsp12:Arg569Gly, Nsp12:Val473Phe, and the associated mutation Nsp12:Met924Leu. As a control we evaluated Nsp12:Phe480Leu and Nsp12:Val557Leu as well (**SI Table 9**). To correct for possible poor protein resolution, we inferred each combination on six different RdRp structures **(SI Figure 5, SI Table 9**).

We detected significant destabilisation only in the case of Nsp12:Met566Val (2.53 kcal/mol) and Nsp12:Val473Phe (3.97 kcal/mol). The former is associated with Nsp12:Met924Leu, which does not show any sort of stabilising effect on its own, nor in combination with any other escape mutation. Nsp12:Val473Phe is strongly associated with a SNP located at genomic position 24378 (*S*:Ser939Phe, adjusted p value=0.00012, **SI Table 7**)

While Ser939Phe yields no effect on pre-fusion S structures (−0.17 kcal/mol), this substitution yields a stabilising effect on post-fusion S structures (−1.9 kcal/mol) **(SI Table 9)**. The physiological gain of this stabilisation is yet to be understood. These results show that escape-associated mutations within RdRp are unlikely to compensate for RdRp stability losses.

## Discussion

The rapid global spread of SARS_CoV-2 has led to the emergence of many different variants in *nsp12*. Our analysis of SARS-CoV-2 RdRp evolution on a global scale demonstrates the negative selection pressure acting on the protein. RdRp mutations potentially linked to remdesivir resistance are rare, and we did not observe consistent impact on protein stability. None of this mutations has yet provided a selective advantage under current pressures. In particular, a Pro323Leu mutation in SARS-CoV-2 genomes from infected patients collected from December 2019 to mid-March 2020 with no reported consequence for RdRp binding site^24^. This finding is in line with the negative selection pressures on *nsp12*, thereby unfavoring resistant variants of SARS-CoV-2. Since mutations in the potential escape motifs of RdRp have been shown to be costly for the related viruses^13^, such variants in SARS-CoV-2 would not be expected to propagate efficiently, unless the virus were constantly challenged by antiviral drug treatment targeting RdRp.

Remdesivir efficacy has been demonstrated previously in SARS-CoV throughout *in vitro* studies of infected cell cultures^25,26,27^, as well as in rhesus macaque models^28^. However, its clinical usage against SARS-CoV-2 has yielded contradictory evidence^14,16^. In the global dataset, the first samples carrying potential remdesivir escape mutations were collected on the 20th of January in China^22^, and 3rd March in the United Kingdom, far before the introduction of remdesivir as a provisionally authorised medicament, (first approval: 1st May, 2020^29^). Those two early mutations did not further spread, as there is no trace in later time points in the global dataset, probably because of purifying selection.

Remdesivir treatment for COVID-19 was initiated at our hospital as a part of a clinical trial in mid March 2020; it was approved for general use in Swiss hospitals as of June 30. Among the patients enrolled in the trial, only eight fulfilled the criteria for treatment with remdesivir in our study period. The only mutation affecting RdRp was not a potential escape motifs, and was found in a non-remdesivir treated patient. Although sampling bias could be the explanation of these results, a potential explanation could be that remdesivir treatment, at the administered doses, does not provide enough selective pressure against the virus due to a low efficacy^30^.

Among the escape mutations showing stable association with other variants, the only instances of destabilisation include Nsp12:Met566Val and Nsp12:Phe480Leu. We found that Nsp12:Met924Leu, the only significantly associated mutation on RdRp, does not rescue Nsp12:Met566Val destabilisation, nor does it have a stabilizing effect on its own. If the recovered escape mutations were costly in terms of fitness, we would have expected to observe a destabilising effect on RdRp, and a correspondent stabilising effect of associated mutations. This is surprisingly not the case, especially considering Nsp12:Arg555Pro, a key residue involved in remdesivir binding, and Nsp12:Val557Leu^18,19^. These results, however, do not exclude that a different form of fitness loss/compensation not involving stability could take place^31^. For example, Nsp12:Arg555Pro could disrupt the binding affinity of remdesivir and Nsp12:Met924Leu could improve the protein binding to the template. In other motifs of RdRp, a depolarising mutation such as Nsp12:Met924Leu has been linked to a reduction of the polymerase proofreading activity^4^. Alternatively such association could involve more widely the viral physiology, as for example the association between the potential escape mutation Nsp12:Val473Phe and S:Ser939Phe. The nature of such cross-protein association remains speculative and additional data would be necessary to illustrate that in full.

The rarity of potential escape mutations and the purifying selection acting on RdRp could indicate that remdesivir use has not yet selected for resistant variants observable on a global scale. However, any selective pressure caused by remdesivir will not necessarily be captured within the entire global dataset (n=56,806), as available sequences represent (i) largely samples from non-remdesivir treated patients (data available from the GISAID repository) and (ii) likely the first isolate from patients, prior to treatment with remdesivir. Therefore, it remains important to continuously sequence isolates from antiviral treated patients and monitor emerging mutations.

## Conclusion

In summary, our study offers a surveillance framework for SARS-CoV-2 evolution focusing on the potential emergence of antiviral resistance. Our findings demonstrate the high conservation of RdRp worldwide and point towards a low selective pressure provided by anti-RdRp drugs (eg. remdesivir). Potential remdesivir escape mutations were very rare, which could be an indicator of little selective pressure and hence no therapeutic effect of remdesivir. Notably, our analysis could be extended to other repurposed RdRp-targeting drugs, such as sofosbuvir and ribavirin, that have the same mechanism of RdRp inhibition as remdesivir.

Our data indicates RdRp as a potential drug target candidate because of the many minor variants screened, yet none of them has gained selective advantage against remdesivir. Patient treated with a potential antiviral drug should be closely monitored for the potential emergence of antiviral resistance.

## Methods

### Inferring signatures of selection in *nsp12*

We inferred selection on the *nsp12* gene using Tajima’s D test statistic, which is the comparison of average number of pairwise differences with the number of segregating sites, as well the ratio of nonsynonymous/synonymous (dN/dS) mutations per codon along *nsp12*. The 58,208 consensus sequences from GISAID that passed COVGAP v2 quality filtering, were further cleaned and filtered from redundant sequences (N = 614) using Sequence Dereplicator and Database Curator (SDDC) program^32^. Since selection analysis tools are limited in the number of sequences that can be handled, we down-sampled to represent all phylogenetic lineages from all countries and each month by 25 sequences using the nextstrain analysis pipeline v.2.0.0 (nextstrain.org)^33^ resulting in 16,184 genomes that could be processed further. Whole-genome consensus sequences were aligned against NCBI Refseq sequence Wuhan-Hu-1 NC_045512.2 using MAFFT v7.467 (method FFT-NS-fragment; options --reorder --keeplength --mapout -- kimura 1 -- addfragments --auto)^34^. The alignment was trimmed to the nucleotide region that codes for *nsp12* (PDB protein identifier 6M71) between 13442-16234, the final stop codon excluded as required for the analysis. Since *nsp12* is translated with a ribosomal slippage, in which the first nine amino acids are read in the second open reading frame (ORF) and the following amino acids in the first ORF, an extra cytosine (C) was included after position 27 to provide the information for all translated codons for subsequent analyses of selection. A last filtering step was applied before the final selection analysis to remove sequences with premature STOP codons (N = 12), remove sequences that contained gaps (−) or missing data (N) (N = 1141), as well as sequences with ambiguous characters (IUPAC codes: YKRWSMDVHB; N = 413). The selection analysis dataset finally contained 14,’612 SARS-CoV-2 consensus sequences.

To infer the selection pressure that is acting on the entire *nsp12* gene region, we calculated Tajimas’s D statistic^35,36^ using MEGA version 7^37^ to test for neutral selection for the entire coding sequence. The test statistic is based on two estimates, number of segregating sites^38^ and average number of nucleotide differences, gained from pairwise comparisons. Tajimas’s D statistics equalling zero means that number of segregating sites roughly equals average nucleotide differences; Tajimas’s D being smaller than zero means that number of segregating sites are more abundant than average nucleotide differences, indicating an excess of rare alleles or a recent population expansion after a bottleneck; and Tajimas’s D being larger than zero means that fewer segregating sites than average nucleotide differences exist, indicating that rare alleles are selected against.

Further, we inferred the ratio of nonsynonymous to synonymous mutations (dN/dS or omega) in a Bayesian sliding window approach using genomegamap^39^. We ran 1,000,000 MCMC chains with 100,000 as burn-ins. The dN/dS ratio per codon is indicative of the kind of selection acting on the codons. Non-synonymous mutations are biologically selected against as they can result in structural changes and hence functional changes unless they provide selective advantage, meaning conveying larger reproductive success. An excess of non-synonymous mutations (dN/dS > 1) is interpreted as the site being under positive selection, whereas an excess of synonymous mutations (dN/dS > 1) is interpreted as purifying or balancing selection acting on this particular position. A balance of non-synonymous and synonymous mutations (dN/dS = 1) is understood as neutral selection. We set the priors of theta (the observed nucleotide diversity of the sample population) to 0.0006 and for kappa (per-path rate bias, from which the transition/transversion ratio can be calculated as kappa/2) to 2.3 as previously inferred in Wilson (2020)^39^.

### Potential escape motifs

Phe480 and Val557 are encoded by nucleotides the codon encompassed between position 14878-14880, and 15109-15111 of SARS-CoV-2 genome (reference NC_045512.2) respectively. We focussed on the motifs neighbouring those two mutations along the following rational: for Val557 we included the entire beta-sheet Val557 finds itself in, and also the loose loops and part of the neighbouring alpha helix closer to the active site^18^. For Phe480, as seemingly no neighbouring structure binds either the template or the nucleoside, we included the entire loose loop and alpha helix neighbouring Phe480, as part of the hydrophobic core of RdRp and crucial for the protein proof-reading stability^4^. The resulting motives stretch from Arg467 to Val493 for the first motif (neighbouring Phe480), and from Leu544 to Gln570 for the second motif (neighbouring Val557).

### Global dataset, spatio-temporal trends and genome entropy

77,150 SARS-CoV-2 consensus genomes were downloaded as of 23rd July 2020 from the GISAID platform^21^. The genomes were quality filtered: 18,942 genomes were discarded as containing more than 10% ambiguous basecalls (Ns); and 1,402 genomes were discarded for containing ambiguous variant calls. Mutations were considered only when a clear alternative allele was found, genomes reporting errors in the alternative alleles calls were discarded. Degenerate basecalls in the variant call other than N were not determining the exclusion of the genome from the dataset. Using our COVGAP pipeline^20^, the sequences were first aligned to the Wuhan-1 reference^40^ using mafft v7.467, point mutations were identified with snp-sites v2.5.1^41^. The collected variants were then annotated using snpeff v4.5covid19^42^. The variants positions, together with the variant annotation, were screened using R base. Genomes whose variants fell into the potential escape motifs were labeled as potential escape mutants.

Statistics on spatio-temporal trends were calculated via R base v3.6.2, lineages classification was inferred through Pangolin^23^. Data mining and structuring was performed through the R packages: tidyr v1.1.2, dplyr v1.0.2, reshape2 v1.4.4^43^, BiocGenerics v0.32.0^44^, IRanges v2.20.2^45^, Biostrings v2.54.0, XVector v0.26.0, S4Vectors v0.24.4, shapefiles v0.7, foreign v0.8-76. Plots were drafted using R packages: ggpubr v0.4.0, ggExtra v0.9, cowplot v1.1.0, lubridate v1.7.9^46^, rgeos v0.5-5, rnaturalearth v0.1.0, rnaturalearthdata v0.1.0, sf v0.9-5, sp v1.4-2, maps v3.3.0, ggspatial v1.1.4, ggplot2 v3.3.2^47^.

Calculation of genome diversity took place using the chao2 index (PMID: 19449706) as available in the R package fossil v0.4.0, the index value was calculated for each single genome, further structuring in table was performed through the Matrix v1.2-18 R package.

### Genome drafting tool and availability

We generated COVGAP2, an adaptation of our bioinformatic COVID-19 Genome Analysis Pipeline (COVGAP, see Online Methods; medRxiv 2020.09.01.20186155; doi:https://doi.org/10.1101/2020.09.01.20186155) to detect all non-synonymous SNPs affecting the defined potential escape motifs and potentially hampering the functionality of drugs targeting RdRp, and to screen sequence data for minor allele variants.

Of note, the COVGAP2 platform can be adapted to detect any relevant variant, including binding sites of new antiviral drugs. This allows us to rapidly screen genome collections and provide molecular epidemiological surveillance in real time or retrospectively.

### Patients, samples, and diagnosis

#### Basel University hospital single time point open cohort

All persons with a first-time positive SARS-CoV-2 PCR test between the 26th of February (first case in Switzerland) and the 30th of April 2020 were eligible for inclusion in the cohort. During this time period, 10,310 diagnostic tests were performed. Of the 690 patients meeting the eligibility criteria, 341 were female and 349 were male, the average age was 48.79 years ±19.64.

#### Longitudinal cohort and remdesivir treatment

All hospitalized SARS-CoV-2 positive patients (diagnosed between 26th of February and 30th of April 2020), with a minimum of two or more positive tests were eligible for inclusion. After exclusion of patients who did not give general consent, the cohort counted 197 patients (114 males, 83 females) with an average age of 62.15 years ±17.66. Patients received 200mg of remdesivir on the first day, 100 mg/day the following days in intravenous infusion for 30 to 60 minutes.

#### Diagnosis and sequencing

The procedure followed to routine-diagnose patients and to sequence the Basel open cohort viral genomes is described in Stange et al., 2020^20^. The region haboring the two potential escape motifs was amplified from the samples from the longitudinal cohort using primers Fwd:AGGAATTACTTGTGTATGCTGCTGA and Rev:TAACATGTTGTGCCAACCACCA, resulting in a 701 bp fragment. The Illumina DNA Prep kit was used to generate sequencing libraries from the fragments libraries before sequencing on an Illumina NextSeq500. This resulted in 259 analyzable samples.

### Statistics and visualisation

#### Genome diversity and statistical test

To calculate the diversity of each genome chao2 index was chosen because if its incidence based nature^48^. Log values were used to ensure normal distribution. Given the large size disparity, a Monte-Carlo approach was chosen. For both first and second motif (n=115, n=69 respectively), the correspondant no-escape mutant group (“None”) (n=56691, n=56737 respectively) was randomly subsampled without replacement in 10000 groups each with n=150. Each subgroup was tested against the same motif group through t-student test. Resulting p-values were adjusted following the Benjamini-Yekutieli correction^49^, and subsequently merged using the sum of logs Fisher method^50^. The R package metap v=1.4 was used.

#### Associated mutation inference, structural fitting and visualisation

To infer escape-associated mutations, all the samples from the global cohort (n=56,806) were labelled based on whether or not they showed at least one non-synonymous mutation in either potential escape motifs, respectively. From the initial 16,051 mutations detected over the entire cohort, only mutations appearing in both non potential escape and potential escape mutants were considered (n=1,056). Generalised linear models were used to evaluate the correlation between each point mutation and the escape/non-escape predictor. Lineages were added as a fixed effect only when the mutation appeared in multiple lineages. The response error was assumed to be distributed binomially. The retrieved p-value was then corrected for multiple testing using Benjamini-Yekutieli correction^49^ and considered significant only for those mutations showing an adjusted p value < 0.05. In order to assess to which response the significant mutations were associated with, we first calculated for each mutation the frequency in escape mutants divided by the frequency in non-escape mutants. We then defined mutations as escape-associated, only if the escape/non-escape frequency ratio was above the 95th percentile of the significant mutation ratio distribution. Conversely, mutations showing a ratio below the 5th percentile were considered not-escape associated. Percentiles were established according to Hyndman and Fan (1996) in order to obtain median-unbiased quantiles. The glm function of R base stats package 3.6.2 was used. The retrieved associated mutations were fitted into the protein structure reported by Yin et al., ^19^ (PBD ID 7BV2). Visualisation and figure drafting took place with pymol version 2.3.5^51^.

### Stability Analysis of Mutation Combinations

The effect of mutations on the stability of RdRp was calculated by using the FoldX 5.0 advanced protein design suite.^52^ FoldX 5.0 has the capabilities to measure the stability changes of a protein structure model upon several mutations and is able to consider the interactions of the protein with RNA in its calculations, which is vital for an analysis in the RdRp system. The input PDB files are identical to those downloaded from the PDB and were not otherwise preprocessed.

We used the FoldX command “BuildModel” and enabled the calculation of interactions between the protein and RNA. For every combination of protein structure and mutations, the calculations of Gibbs energies of protein folding were repeated five times, and the differences between respective wild types and mutated proteins were reported. We then calculated the average total Gibbs energy difference over the six protein structures of RdRp to have a final estimate of the stability difference between the wild type and the mutated protein.

To distinguish genuine stability predictions from noise, we filtered the results based on the reported standard deviations of FoldX energy calculations as described in Buss et al^53^. The accuracy of FoldX is described to be dependent on the resolution of the investigated structures. Given that the resolution of the used protein structures ranges on the lower end with values between 2.5 and 2.9 Ångstrom, we used the conservative threshold of 1.78 kcal/mol.

Energy differences below this threshold were not considered to denote changes in the stability of the investigated protein.

To have a diverse perspective on the impact of the candidate resistance and compensatory mutations, a stability analysis was conducted on multiple experimentally resolved structures of the SARS-CoV-2 RdRp: 7CXM, 7AAP, 7BV2, 7C2K, 6YYT, and 6M71. The PDB files of these structures were directly downloaded from the Protein Data Bank^54^.

## Supporting information

GISAID_acknowledgements_0

GISAID_acknowledgements_1

GISAID_acknowledgements_2

GISAID_acknowledgements_3

GISAID_acknowledgements_4

GISAID_acknowledgements_5

GISAID_acknowledgements_6

GISAID_acknowledgements_7

## Data Availability

Genome assemblies from the Basel open cohort were submitted to GISAID. Amplicon sequences from the remdesivir treated patients are available upon request.

## Ethics

The study was conducted according to good laboratory practice and in accordance with the Declaration of Helsinki and national and institutional standards and was approved by the ethical committee (EKNZ 2020-00769). The clinical trial accession numbers are NCT04323761 and NCT04351503 (clinicaltrials.gov).

## Authors contribution

AE and HH devised the study. AlM designed COVGAP2, drafted the genomes of the cohorts, mined the datasets and the models, drafted the manuscript. TR and HSS supervised the sequencing and the sample collection, deposited genomic data to GISAID. MaSt performed the selection pressure analysis on *nsp12*. KKS and MiSc collected clinical information on the inpatient cohort. EA, GT, LTA drafted the stability analysis upon mutations of RdRp. MiSc, KL, AG collaborated in sample collection. AuM, JB, HP, MaSi, CN, RB, MO, SB, SP, MB, CM, TS, provided clinical expertise and valuable discussion on the results.

## Acknowledgements

We are grateful to Dr. Robert Ivanek, Dr. Julien Roux and Dr. Florian Geier (all Department of Biomedicine (DBM) Bioinformatics Core Facility, University of Basel) for the consultancy provided on the generalised linear models, as well as on the selection pressure analysis. We thank the clinical virology team (University Hospital Basel), in particular Nadine Blind for their efforts to gather and extract samples. Furthermore, we thank Daniel Gander, Christine Kiessling, Magdalena Schneider, Elisabeth Schultheiss, Clarisse Straub, and Rosa-Maria Vesco (University Hospital Basel) for excellent technical assistance with next generation sequencing. Calculations were performed at sciCORE (http://scicore.unibas.ch/) scientific computing center at University of Basel, the support from the sciCORE team for the analysis is greatly appreciated. Support for the creation of schematic figures (S1) was provided by BioRender.com. We thank all authors who have shared their genomic data on GISAID, particularly to the group of Prof. Dr. Tanja Stadler from ETH Zürich for the submission of large part of the Swiss genomes. A full table outlining the originating and submitting labs is included as a supplementary file.

## Supplementary data

**Figure S1:**
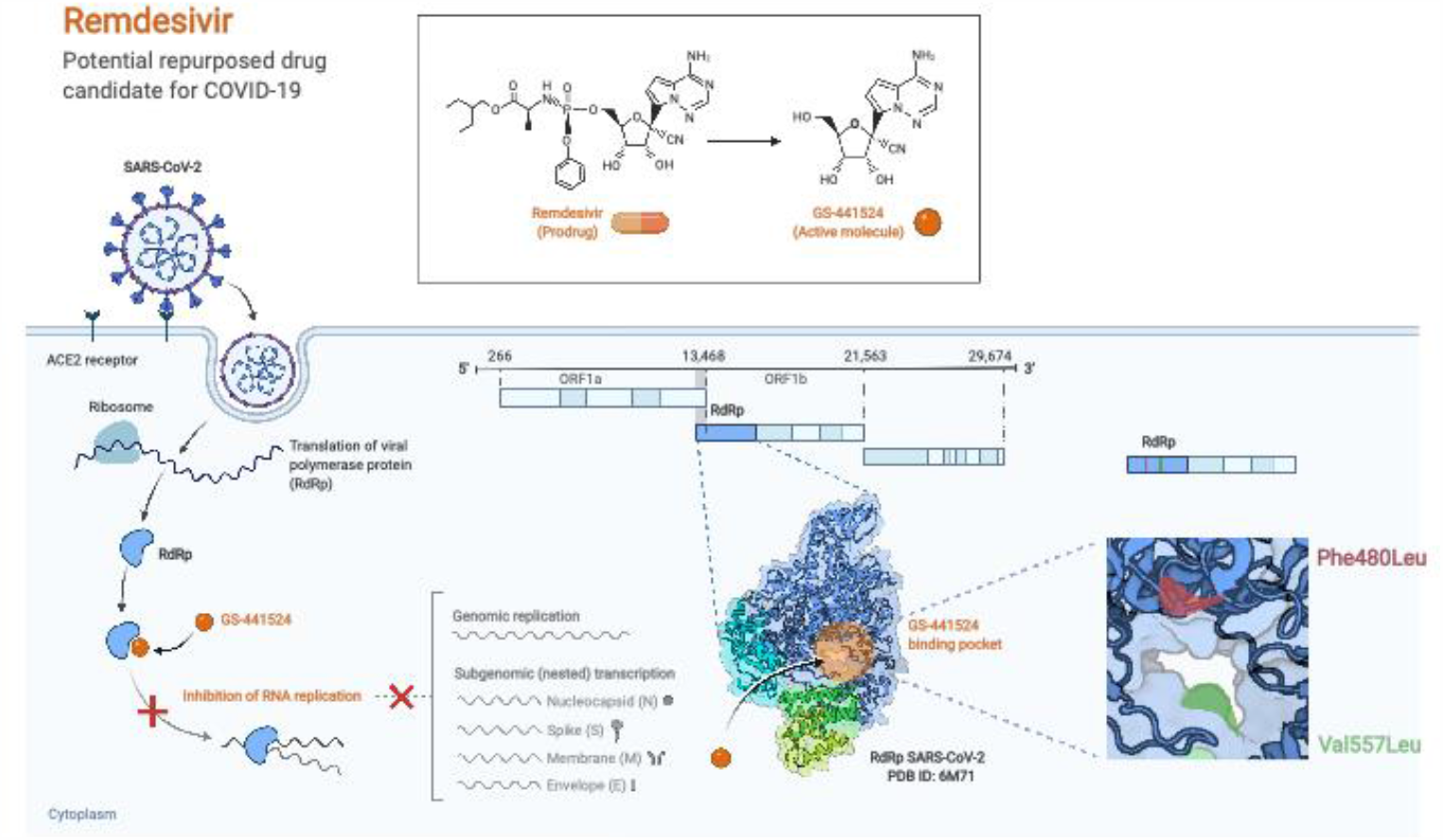
Remdesivir mechanism of action. The prodrug is metabolized into a ATP analogue, and triggers the termination of RNA polymerization after binding to the active site. the two mutation are schematized in their location, Phe480 buried, Val557 adjacent to the binding site. Credits to Biorender.com

**Figure S2:**
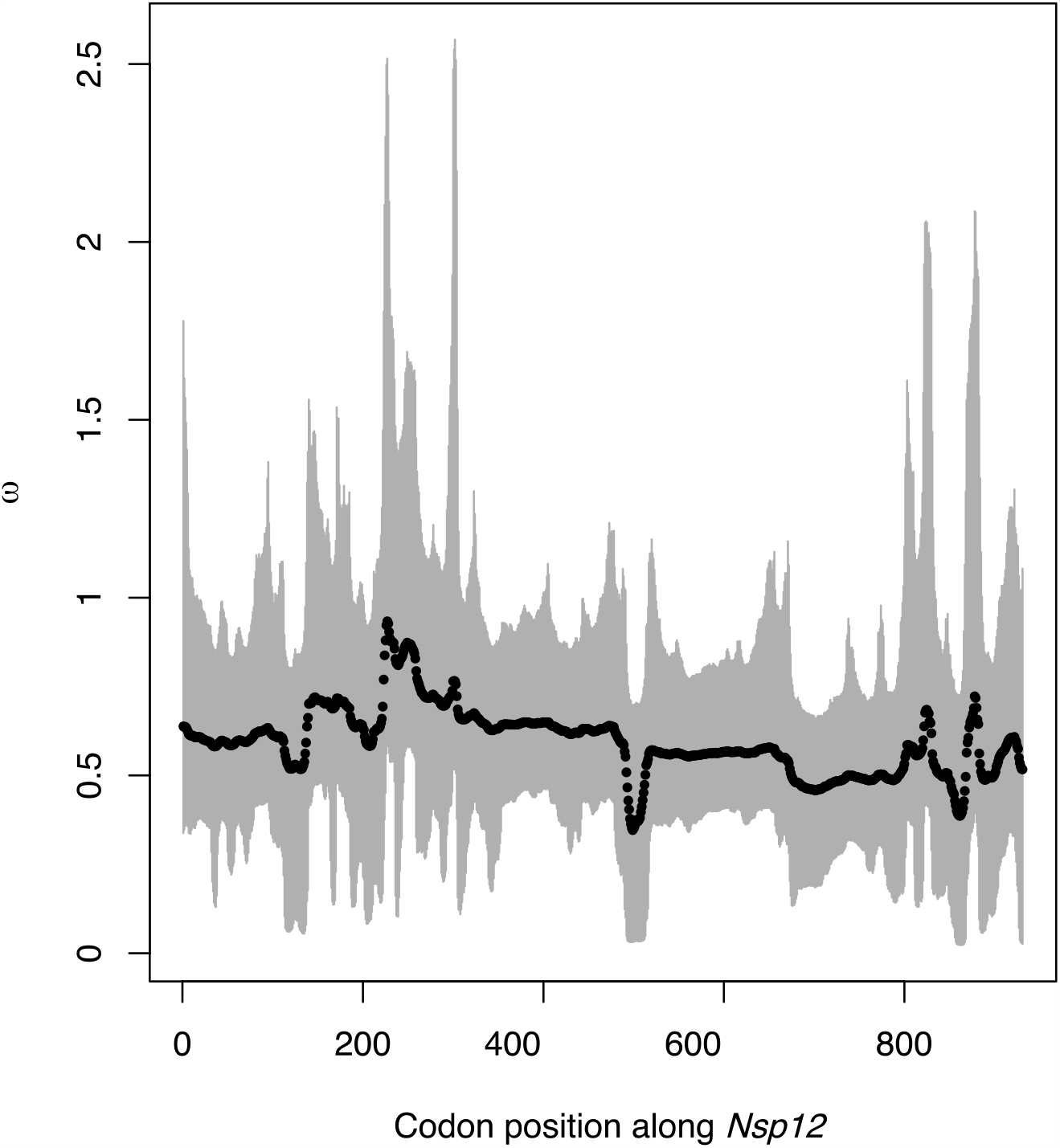
Codon-based estimates of the ratio of nonsynonymous to synonymous mutations (omega, dN/dS) along *nsp12* using a sliding window model in a Bayesian computation approach. An excess of non-synonymous mutations (dN/dS > 1) is interpreted as the site being under positive selection, whereas an excess of synonymous mutations (dN/dS < 1) is interpreted as purifying or balancing selection acting on this particular position. A balance of non-synonymous and synonymous mutations (dN/dS = 1) is understood as neutral selection.

**Figure S3:**
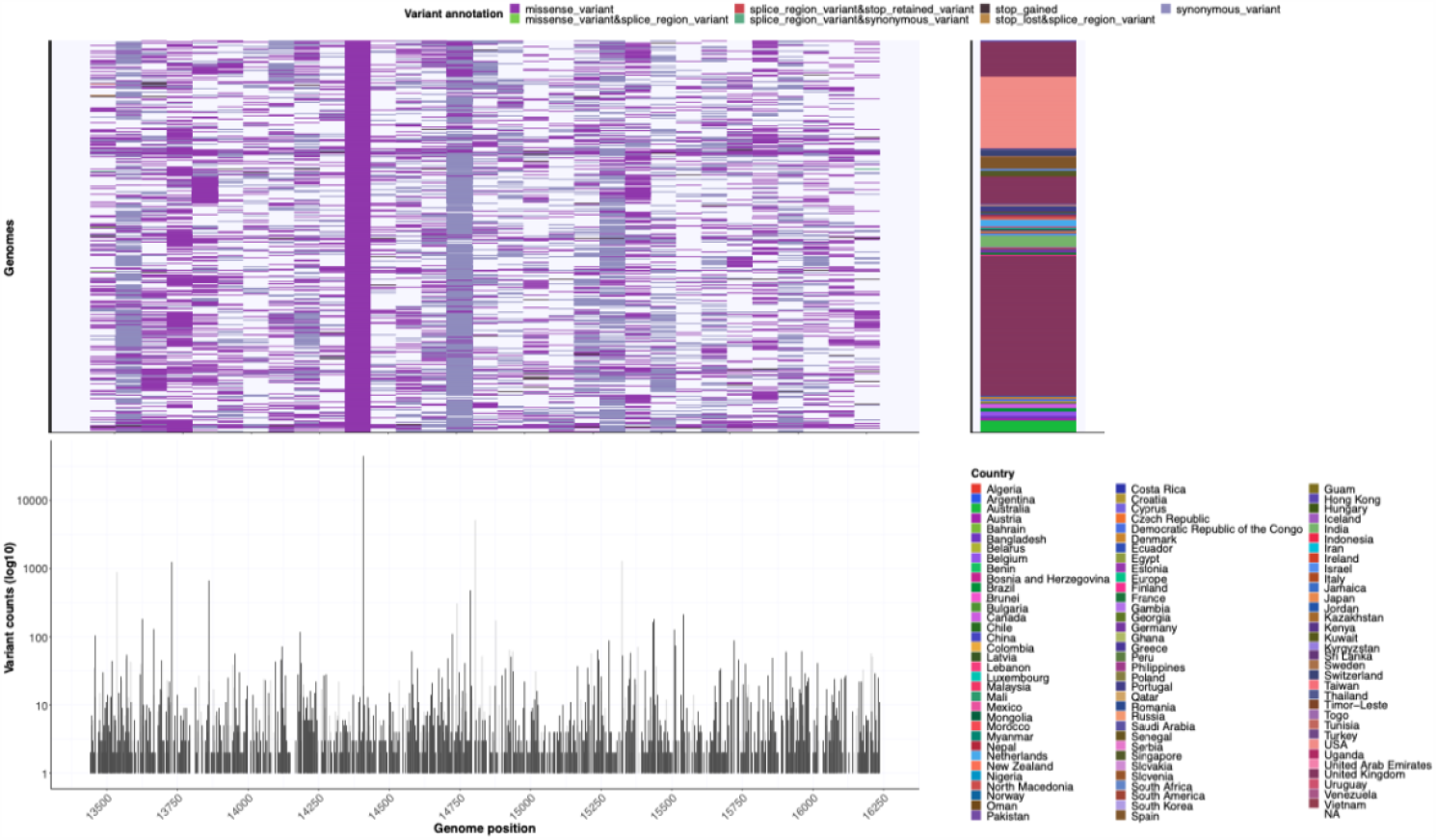
Mutation distribution in Rdrp shows hotspots of variants mostly not affecting the binding site of template and remdesivir. The top right displays mutation prevalence in RdRp, bottom left the frequencies, top left the originating countries

**Figure S4:**
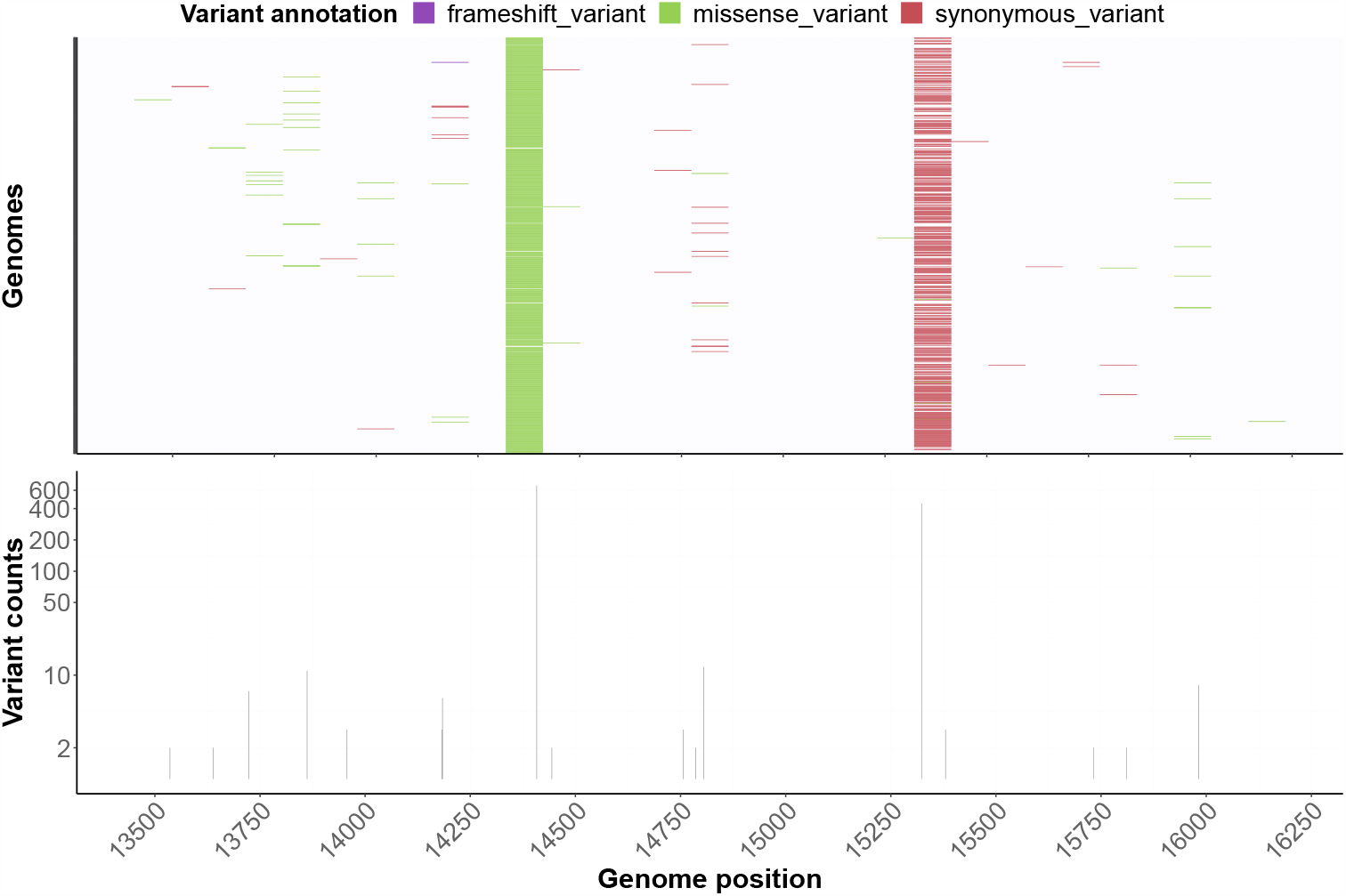
Basel open cohort: mutation distribution in RdRp is in line with the global trend, except for a mutation with high prevalence in the larger Basel are. Depiction of mutation in single-timepoint tested patients in University Hospital Basel (n=690) from 23rd February 2020 till 30th April 2020. Upper panel illustrates mutation prevalence, bottom panel the square-rooted frequency

**Figure S5:**
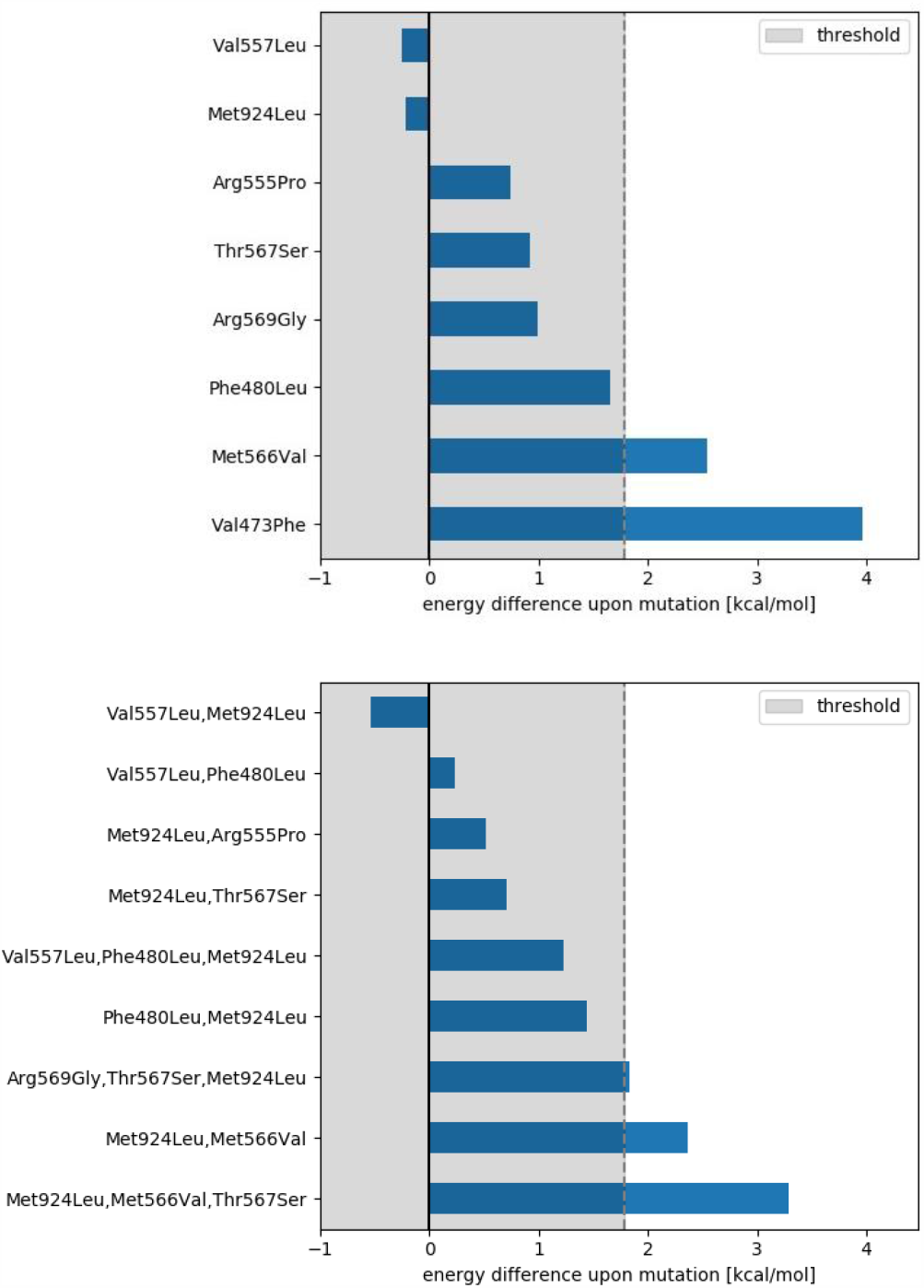
Average energy difference between wild-type and mutated RdRp across six different experimental structures. Destabilizing mutations have a positive energy difference while stabilizing mutations are negative. The panel above shows the single variants while the panel below depicts the found mutation combinations. The threshold upon which an energy difference is considered significant is marked with a light grey rectangle.

## References

1. Liu, X. & Wang, X.-J. Potential inhibitors against 2019-nCoV coronavirus M protease from clinically approved medicines. J. Genet. Genomics 47, 119–121 (2020).

2. Coleman, C. M. et al. Abelson Kinase Inhibitors Are Potent Inhibitors of Severe Acute Respiratory Syndrome Coronavirus and Middle East Respiratory Syndrome Coronavirus Fusion. J. Virol. 90, 8924–8933 (2016).

3. Stader, F. et al. Stopping lopinavir/ritonavir in COVID-19 patients: duration of the drug interacting effect. J. Antimicrob. Chemother. 75, 3084–3086 (2020).

4. Shannon, A. et al. Remdesivir and SARS-CoV-2: Structural requirements at both nsp12 RdRp and nsp14 Exonuclease active-sites. Antiviral Res. 178, 104793 (2020).

5. Selisko, B., Papageorgiou, N., Ferron, F. & Canard, B. Structural and Functional Basis of the Fidelity of Nucleotide Selection by Flavivirus RNA-Dependent RNA Polymerases. Viruses 10, (2018).

6. Elfiky, A. A. Ribavirin, Remdesivir, Sofosbuvir, Galidesivir, and Tenofovir against SARS-CoV-2 RNA dependent RNA polymerase (RdRp): A molecular docking study. Life Sci. 253, 117592 (2020).

7. Chien, M. et al. Nucleotide Analogues as Inhibitors of SARS-CoV-2 Polymerase, a Key Drug Target for COVID-19. J. Proteome Res. 19, 4690–4697 (2020).

8. Jockusch, S. et al. Sofosbuvir terminated RNA is more resistant to SARS-CoV-2 proofreader than RNA terminated by Remdesivir. Sci. Rep. 10, 16577 (2020).

9. Bartholomeusz, A., Tehan, B. G. & Chalmers, D. K. Comparisons of the HBV and HIV polymerase, and antiviral resistance mutations. Antivir. Ther. 9, 149–160 (2004).

10. Eltahla, A. A., Luciani, F., White, P. A., Lloyd, A. R. & Bull, R. A. Inhibitors of the Hepatitis C Virus Polymerase; Mode of Action and Resistance. Viruses 7, 5206–5224 (2015).

11. Bloom, J. D., Gong, L. I. & Baltimore, D. Permissive secondary mutations enable the evolution of influenza oseltamivir resistance. Science 328, 1272–1275 (2010).

12. Ali, S. et al. Selected replicon variants with low-level in vitro resistance to the hepatitis C virus NS5B polymerase inhibitor PSI-6130 lack cross-resistance with R1479. Antimicrob. Agents Chemother. 52, 4356–4369 (2008).

13. Agostini, M. L. et al. Coronavirus Susceptibility to the Antiviral Remdesivir (GS-5734) Is Mediated by the Viral Polymerase and the Proofreading Exoribonuclease. mBio 9, e00221–18 (2018).

14. Beigel, J. H. et al. Remdesivir for the Treatment of Covid-19 - Final Report. N. Engl. J. Med. 383, 1813–1826 (2020).

15. Spinner, C. D. et al. Effect of Remdesivir vs Standard Care on Clinical Status at 11 Days in Patients With Moderate COVID-19: A Randomized Clinical Trial. JAMA 324, 1048–1057 (2020).

16. WHO Solidarity Trial Consortium et al. Repurposed Antiviral Drugs for Covid-19 - Interim WHO Solidarity Trial Results. N. Engl. J. Med. (2020) doi:10.1056/NEJMoa2023184.

17. Wang, Y. et al. Remdesivir in adults with severe COVID-19: a randomised, double-blind, placebo-controlled, multicentre trial. Lancet Lond. Engl. 395, 1569–1578 (2020).

18. Gao, Y. et al. Structure of the RNA-dependent RNA polymerase from COVID-19 virus. Science 368, 779–782 (2020).

19. Yin, W. et al. Structural basis for inhibition of the RNA-dependent RNA polymerase from SARS-CoV-2 by remdesivir. Science 368, 1499–1504 (2020).

20. Stange, M. et al. SARS-CoV-2 outbreak in a tri-national urban area is dominated by a B.1 lineage variant linked to mass gathering events. medRxiv 2020.09.01.20186155 (2020) doi:10.1101/2020.09.01.20186155.

21. Elbe, S. & Buckland-Merrett, G. Data, disease and diplomacy: GISAID’s innovative contribution to global health. Glob. Chall. 1, 33–46 (2017).

22. Jiang, X.-L. et al. Transmission Potential of Asymptomatic and Paucisymptomatic Severe Acute Respiratory Syndrome Coronavirus 2 Infections: A 3-Family Cluster Study in China. J. Infect. Dis. 221, 1948–1952 (2020).

23. Rambaut, A. et al. A dynamic nomenclature proposal for SARS-CoV-2 lineages to assist genomic epidemiology. Nat. Microbiol. 5, 1403–1407 (2020).

24. Pachetti, M. et al. Emerging SARS-CoV-2 mutation hot spots include a novel RNA-dependent-RNA polymerase variant. J. Transl. Med. 18, 179 (2020).

25. Wang, M. et al. Remdesivir and chloroquine effectively inhibit the recently emerged novel coronavirus (2019-nCoV) in vitro. Cell Res. 30, 269–271 (2020).

26. Gordon, C. J. et al. Remdesivir is a direct-acting antiviral that inhibits RNA-dependent RNA polymerase from severe acute respiratory syndrome coronavirus 2 with high potency. J. Biol. Chem. 295, 6785–6797 (2020).

27. Pruijssers, A. J. et al. Remdesivir Inhibits SARS-CoV-2 in Human Lung Cells and Chimeric SARS-CoV Expressing the SARS-CoV-2 RNA Polymerase in Mice. Cell Rep. 32, 107940 (2020).

28. Williamson, B. N. et al. Clinical benefit of remdesivir in rhesus macaques infected with SARS-CoV-2. Nature 585, 273–276 (2020).

29. Lamb, Y. N. Remdesivir: First Approval. Drugs 80, 1355–1363 (2020).

30. Götte, M. The distinct contributions of fitness and genetic barrier to the development of antiviral drug resistance. Curr. Opin. Virol. 2, 644–650 (2012).

31. Storz, J. F. Compensatory mutations and epistasis for protein function. Curr. Opin. Struct. Biol. 50, 18–25 (2018).

32. Ibrahim, E. S., Kashef, M. T., Essam, T. M. & Ramadan, M. A. A Degradome-Based Polymerase Chain Reaction to Resolve the Potential of Environmental Samples for 2,4-Dichlorophenol Biodegradation. Curr. Microbiol. 74, 1365–1372 (2017).

33. Hadfield, J. et al. Nextstrain: real-time tracking of pathogen evolution. Bioinforma. Oxf. Engl. 34, 4121–4123 (2018).

34. Katoh, K. & Standley, D. M. MAFFT Multiple Sequence Alignment Software Version 7: Improvements in Performance and Usability. Mol. Biol. Evol. 30, 772–780 (2013).

35. Tajima, F. Statistical method for testing the neutral mutation hypothesis by DNA polymorphism. Genetics 123, 585–595 (1989).

36. Nei, M. & Kumar, S. Molecular Evolution and Phylogenetics. (Oxford University Press, 2000).

37. Kumar, S., Stecher, G. & Tamura, K. MEGA7: Molecular Evolutionary Genetics Analysis Version 7.0 for Bigger Datasets. Mol. Biol. Evol. 33, 1870–1874 (2016).

38. Watterson, G. A. On the number of segregating sites in genetical models without recombination. Theor. Popul. Biol. 7, 256–276 (1975).

39. Wilson, D. J. & CRyPTIC Consortium. GenomegaMap: Within-Species Genome-Wide dN/dS Estimation from over 10,000 Genomes. Mol. Biol. Evol. 37, 2450–2460 (2020).

40. Wu, F. et al. A new coronavirus associated with human respiratory disease in China. Nature 579, 265–269 (2020).

41. Page, A. J. et al. SNP-sites: rapid efficient extraction of SNPs from multi-FASTA alignments. Microb. Genomics 2, e000056 (2016).

42. Cingolani, P. et al. A program for annotating and predicting the effects of single nucleotide polymorphisms, SnpEff: SNPs in the genome of Drosophila melanogaster strain w1118; iso-2; iso-3. Fly (Austin) 6, 80–92 (2012).

43. Wickham, H. Reshaping Data with the reshape Package. J. Stat. Softw. 21, 1–20 (2007).

44. Huber, W. et al. Orchestrating high-throughput genomic analysis with Bioconductor. Nat. Methods 12, 115–121 (2015).

45. Lawrence, M. et al. Software for computing and annotating genomic ranges. PLoS Comput. Biol. 9, e1003118 (2013).

46. Grolemund, G. & Wickham, H. Dates and Times Made Easy with lubridate. J. Stat. Softw. 40, 1–25 (2011).

47. Wickham, H., Chang, W. & RStudio. ggplot2: Create Elegant Data Visualisations Using the Grammar of Graphics. (2016).

48. Chao, A. Estimating the Population Size for Capture-Recapture Data with Unequal Catchability. Biometrics 43, 783–791 (1987).

49. Benjamini, Y. & Yekutieli, D. The Control of the False Discovery Rate in Multiple Testing under Dependency. Ann. Stat. 29, 1165–1188 (2001).

50. Fisher, R. A. Statistical Methods for Research Workers. in Breakthroughs in Statistics: Methodology and Distribution (eds. Kotz, S. & Johnson, N. L.) 66–70 (Springer, 1992). doi:10.1007/978-1-4612-4380-9_6.

51. Lill, M. A. & Danielson, M. L. Computer-aided drug design platform using PyMOL. J. Comput. Aided Mol. Des. 25, 13–19 (2011).

52. Schymkowitz, J. et al. The FoldX web server: an online force field. Nucleic Acids Res. 33, W382–388 (2005).

53. Buß, O., Rudat, J. & Ochsenreither, K. FoldX as Protein Engineering Tool: Better Than Random Based Approaches? Comput. Struct. Biotechnol. J. 16, 25–33 (2018).

54. Berman, H. M. et al. The Protein Data Bank. Nucleic Acids Res. 28, 235–242 (2000).

